# Investigating the use and impact of community Care (Education) and Treatment Reviews (C(E)TRs) in people with intellectual disability and autistic people: protocol for a cohort study using electronic health records

**DOI:** 10.1101/2025.07.09.25331199

**Authors:** Brónagh McCoy, Lauren Bell, Kang Wang, Huajie Jin, Angela Hassiotis, André Strydom, Johnny Downs, Ben Carter, Hitesh Shetty, Robert Stewart, Afia Ali, Rory Sheehan

## Abstract

Investigating the use and impact of community Care (Education) and Treatment Reviews (C(E)TRs) in people with intellectual disability and autistic people: protocol for a cohort study using electronic health records

**Introduction:** Care (Education) and Treatment Reviews (C(E)TRs) are intended to reduce unnecessary psychiatric hospital admission and length of stay for people with intellectual disability and autistic people. The use and impact of C(E)TRs has not been systematically evaluated since their introduction in England in 2015. The aims of this study are to describe the demographic and clinical profiles of people who receive a community C(E)TR and to investigate their effects on admission, length of hospital stay, and clinical and functional change.

**Methods and analysis:** We will conduct a retrospective cohort study using de-identified data from electronic health records derived from two large National Health Service mental health providers in London, England, including one replication site. Data will be extracted using the Clinical Record Interactive Search (CRIS) tool for all people with recorded intellectual disability and/or autism who received mental healthcare from 2015. We will identify community C(E)TR events using keyword searches. Community C(E)TRs will be examined in two ways: 1) in a community cohort, we will capture data in the 6-month periods before and after a community C(E)TR and compare this to a matched control group, and 2) in a hospital cohort, we will compare groups who did and did not receive a community C(E)TR prior to their admission. We will describe the socio-demographic and clinical profiles of each group and their health service use and compare C(E)TR and no C(E)TR groups using t-tests (or a non-parametric equivalent). The primary outcomes are admission to psychiatric hospital (community cohort) and length of psychiatric hospital admission and clinical change (hospital cohort), which will be estimated using propensity score weighting and difference-in-differences methods for psychiatric hospitalization, and Cox’s proportional hazard model for length of hospital admission, and repeated-measures ANOVA for clinical change.

**Ethics and dissemination:** Use of CRIS to examine de-identified clinical data for research purposes has overarching ethical approval. This study has been granted local approval by the South London and Maudsley CRIS Oversight Committee. Findings will be disseminated in an open access peer-reviewed academic publication, at conference presentations, and to service users and carers in accessible formats.

**Strengths and limitations of this study:**

- This study will provide a much-needed evidence base to guide the provision of community C(E)TRs their effectiveness in the care pathway of people with intellectual disability and autistic people.
- Routinely collected health data represent real-world clinical practice and enable us to generate a relatively large cohort not biased by participant selection.
- The study will be undertaken in a large NHS Trust with a diverse catchment area. We will replicate the study in a second NHS Trust to provide evidence for the generalizability of the initial findings across a different population.
- The number of people in the dataset who have received a community C(E)TRs is unknown; if there are relatively few events, our dataset may be under-powered to demonstrate differences between those who do and do not receive the intervention (type 2 error).
- Electronic health record data are generated as part of routine clinical practice. There may be a high level of missing data which could limit our analysis.

## INTRODUCTION

Reducing unnecessary psychiatric hospital admissions and length of hospital stay for people with intellectual disability and autistic people has been a focus of English national health policy since the scandal of abuse and neglect of patients at Winterbourne View Hospital in 2011.^1^ The Transforming Care programme was established to improve community health and social care for people with intellectual disability and autistic people and to reduce what had generally been considered to be an over-reliance on in-patient psychiatric services.^2^ A key element of Transforming Care has been the introduction of Care (Education) and Treatment Reviews (C(E)TRs), person-centred review meetings that aim to ensure the right support is provided.^3^ A community C(E)TR should be convened when a person with intellectual disability or an autistic person living in the community is considered at high risk of admission to a psychiatric hospital, either because of mental ill-health or because of an escalation in behaviours that challenge.

C(E)TRs are chaired by a health commissioner and includes an independent clinical expert and expert-by-experience, often a family carer of a person with intellectual disability or autistic person. This panel gathers information from the individual, their families and carers, and professionals, and produces a report and action plan following the C(E)TR which includes recommendations that can enable the person to remain living safely in the community (e.g. input from specialist professionals or health teams, provision of additional care, use of respite services). Where admission to hospital is considered necessary, the C(E)TR panel will define the purpose of the admission and encourage early discharge planning in order to minimise the length of the hospital stay. In-patient C(E)TRs are convened at various points of a hospital admission; the current project investigates only C(E)TRs that are held in the community.

Little formal research has been carried out on community C(E)TRs, such as who receives them and when, and how C(E)TRs may influence health service use, including admission to hospital. By understanding the demographic and clinical profile of patients who do, and not, have a community C(E)TR, it may be possible to develop targeted interventions to improve access to community C(E)TRs and ensure the potential benefits are available to everyone.

### Study aims and design

We will conduct a retrospective cohort study using a large dataset of real-world clinical data derived from Electronic Health Records (EHRs) to investigate who receives a community C(E)TR and the association of community C(E)TRs with patient outcomes and service use. This work includes two sub-studies:

1) Sub-study 1: Community cohort

We will: i) examine the demographic and clinical profile of those who receive a community C(E)TR, and ii) attempt to model the causal relationship of receiving a community C(E)TR on the risk of psychiatric hospital admission.

2) Sub-study 2: Hospital cohort

For patients who are admitted to psychiatric hospital, we will compare: i) the demographic and clinical profile before admission, ii) length of stay in hospital, and iii) clinical and functional improvement whilst in hospital, between those who had a community C(E)TR prior to their admission and those who did not.

## METHODS AND ANALYSIS

### Study setting

Data will be obtained from the South London and Maudsley (SLaM) NHS Foundation Trust. SLaM is one of the largest providers of secondary mental healthcare in Europe with a diverse catchment area of approximately 1.3 million people who live in the south London boroughs of Lambeth, Southwark, Lewisham and Croydon. Mental health services provided by SLaM include specialist teams for children and adolescents with mental illness and/or neurodevelopmental conditions, community teams for adults, including specialist teams for those with intellectual disability, and mainstream and specialist in-patient wards across different sites. The Trust has implemented a full EHR since 2006 and the SLaM NIHR Maudsley Biomedical Research Centre (BRC) Case Register contains the clinical records of >500,000 patients.^4,5^ Unless otherwise stated, data will be extracted up to an end date of 31 December 2024.

### Data source and linkage

The Clinical Record Interactive Search (CRIS) is a research tool that enables analysis of de-identified EHRs. Patient-level data are extracted from structured fields and through interrogation of unstructured free text (e.g. clinical letters and reports) using validated Natural Language Processing (NLP) applications. Available variables include socio-demographic information, neuro-psychiatric signs and symptoms, interventions (medication, psychological therapy), hospital admissions, and contacts with professionals and teams (e.g. doctors, allied health professionals). We will link data obtained from the SLaM clinical record with Hospital Episode Statistics (HES), a national dataset curated by NHS Digital that includes details of all general hospital in-patient admissions, out-patient appointments and attendances to A&E in England.^6^ Data linkage takes place in a secure data safe haven.^7^

### Natural Language Processing

NLP is an information extraction technique used to identify relevant information from unstructured free text via automated methods.^8^ NLP models are developed by defining a construct of interest (e.g. aggression) and manually annotating examples in a set of training data to classify true positive instances of the construct. Various clinically-relevant NLP applications have been developed for use in the SLaM EHR.^9^ NLP-derived data in this study will be obtained as binary variables which indicate the presence of at least one mention of the construct of interest within a given surveillance period.

### Study eligibility criteria

Individuals of any age with a diagnosis of intellectual disability (ICD-10 code F7x) or autism (ICD-10 code F84) will be identified using diagnostic information entered into structured fields or in the free text record using the NLP diagnosis application. Participants must have a recorded diagnosis of intellectual disability and/or autism. Eligible participants must have received an episode of care from a community mental health service provided by SLaM for at least one continuous year between 01 January 2015 and 31 December 2023 to ensure that sufficient data are held in the record. All eligible episodes of care will be included, including those individuals with more than one period of community care of at least one year during this timeframe.

### Identification of C(E)TRs

We will undertake keyword searches in the EHR of people with intellectual disability or autism who meet the study eligibility criteria to identify each community C(E)TR event and the date on which it occurred. Search terms will be “Care and Treatment Review,” “Care Education and Treatment Review,” “CTR,” and “CETR.” The search interface does not recognise punctuation or wildcards. We will create a list of people whose health records contain any of these text strings and will review the context of the positive mention to ascertain true instances of C(E)TR meetings along with the date that the meeting was held. We will label a C(E)TR meeting as having taken place where there is evidence that a meeting has occurred and where a date of the meeting is available. We expect there will be mentions of the search terms which do not correspond to a C(E)TR meeting taking place e.g. where a possible future C(E)TR is flagged. Some mentions of the search terms will be false positives (e.g. spe*ctr*um, ele*ctr*ocardiogram), and such irrelevant cases will be discounted.

To ensure reliability of identification, two members of the research team will independently review records of 100 eligible patients where C(E)TR search terms have been identified. Cohen’s Kappa will be calculated to test agreement between the two raters.^10^ If inter-rater reliability is acceptable (Cohen’s kappa>0.6), the raters will work independently and only flag cases that cannot easily be determined. If initial inter-rater reliability is inadequate (Cohen’s kappa<0.6), we will discuss and refine the method of ascertaining whether a key word mention represents a C(E)TR meeting using a series of test cases. The raters will reconvene periodically to double-review a subset of records to ensure that practice remains consistent. We will retain key excerpts of anonymised text from the EHR as exemplars of mentions of C(E)TRs; a selection of these will be reported for transparency, and the examples can also be used to train any additional members of the research team who interrogate the records.

### Classification of C(E)TRs as community or in-patient

Information on all psychiatric in-patient spells will be extracted for those in the cohort. We will place this alongside the dates of recorded C(E)TRs to categorise the C(E)TR as a community C(E)TR (i.e. occurring when the person was living in the community) or an in-patient C(E)TR (i.e. occurring when the person was admitted to a psychiatric hospital). All analyses that follow will be performed on community C(E)TRs.

### Sub-study 1: Community cohort

**PICO framework** (Table 1)

**Table 1.** PICO framework for sub-studies 1 and 2.

|  | **Sub-Study 1** | **Sub-Study 2** |
| --- | --- | --- |
| **Population** | People of any age with a recorded diagnosis of intellectual disability (ICD-10 code F7x), autism (ICD-10 code F84), or both, with at least one | People of any age with a recorded diagnosis of intellectual disability (ICD-10 code F7x), autism (ICD-10 code F84), or both, with at least one year of care under secondary |
|  | year of care under secondary mental healthcare services. | mental healthcare services and an admission to a psychiatric in-patient ward for a period of at least 14 days. |
| **Intervention** | Community C(E)TR | Community C(E)TR within 6 months prior to admission |
| **Comparison** | No community C(E)TR | Did not receive a community C(E)TR within 6 months prior to admission |
| **Outcome** |  |  |
| *Primary* | Psychiatric hospital inpatient admission within 6 months of the community C(E)TR | Length of psychiatric hospital admission (days) |
| *Secondary* | Descriptive data and comparison of baseline demographics between intervention and comparison groups<br>Descriptive data of 6 months pre-/post-C(E)TR (index date): neuropsychiatric symptoms, clinical variables, and service use | Descriptive data and comparison of baseline demographics between intervention and comparison groups<br>Descriptive data and comparison of pre-hospital admission neuropsychiatric symptoms, HoNOS, and service use<br>Clinical and social functional improvement (change in HoNOS) whilst in hospital |

C(E)TR, Care (Education) and Treatment Review; HoNOS, Health of the Nation Outcome Scales

#### Index date

The index date will be defined as the date on which a community C(E)TR was held.

#### Exposure

Exposure will be defined as the community C(E)TR.

#### Descriptive Measures

Baseline measures extracted at the index date will include age, sex, ethnicity, deprivation score (Index of Multiple Deprivation^11^), and other socio-demographic, mental and physical health indicators (Supplementary Tables 1 and 2).

#### Outcome

The primary outcome will be psychiatric hospital admission (treated as a binary variable) at any time within 6 months after the index date.

### Matching a control group

We will match between one and five controls, who will not have a community C(E)TR, per patient who has a C(E)TR. Eligibility will be a diagnosis of intellectual disability or autism and having received a continuous period of community care within SLaM services of at least a year between 1 January 2015 and 31 December 2023. Matching will be based on age (+/-3 years), sex (male/female), and diagnosis (intellectual disability with/without autism, or autism without intellectual disability). Controls should be under an active episode of community care and not be an inpatient on the index date of their matched case. They must also have been under SLaM services for a similar total duration of their matched case (+/- 2 years). Matching with replacement will be applied to ensure every case has an appropriate match according to these criteria. A pseudo-index date for each control will be assigned and will be the same as the index date of their matched case.

### Multiple C(E)TRs

Community C(E)TRs of greater than six months apart for the same patient will be treated as separate events. As community C(E)TRs require substantial planning and organisation, we expect few to take place within six months of a previous C(E)TR; however, if this occurs, the earlier C(E)TR date will be used as the index date and a second C(E)TR within the six month timeframe will be excluded from analysis.

### Statistical analysis

#### Descriptive statistics

We will report baseline socio-demographic variables for the C(E)TR and control groups (Supplementary Tables 1 and 2) Frequencies and percentages will be reported for categorical variables and mean (with standard deviation) or median (with inter-quartile range) for continuous and count variables, depending on their distributions. We will describe these data separately for two diagnostic categories: 1) people with intellectual disability with/without autism, and 2) autistic people without an intellectual disability. We will compare the C(E)TR to the control group on these baseline socio-demographic characteristics using standard statistical comparisons for categorical (Chi square) and continuous variables (t-test or Kruskall-Wallis).

Descriptives of clinical variables and statistical comparisons between the C(E)TR and control groups will be provided separately for the 6-month period before the index date and the 6-month period after the index date. These descriptives will not lead to a causal interpretation of the effect of C(E)TRs.

#### Causal effect of community C(E)TRs on risk of hospitalization

A two-stage framework will be applied to evaluate the causal effect of having a community C(E)TR on hospital in-patient prevalence after the C(E)TR; propensity score weighting (propensity model) and a difference-in-differences (DiD) analysis (outcome model). We will compare changes from the 6-month period before the index date and the 6-month period after the index date, between people with intellectual disability and autistic people who had a C(E)TR and those who did not. The outcome will be whether a person has a psychiatric in-patient admission at any time in the 6 months after the index date.

#### Stage 1: Propensity scores

As the retrospective analysis of EHRs is not a randomized controlled study, we will employ a propensity score weighting technique to adjust for confounders in the data. Propensity scores will balance the C(E)TR and comparison groups on a set of baseline (pre-index date) characteristics.^12^ Certain characteristics may increase the probability of someone receiving a C(E)TR and we wish to account for these in the outcome (risk of hospitalization) model. Scores will be calculated by fitting a multivariable logistic regression model with C(E)TR status as the dependent variable.^13^ We will condition on baseline characteristics that are believed to be related to the outcome, risk of hospitalization, and to the probability of receiving a C(E)TR (confounders), but not those that might lie on the causal pathway from receiving a C(E)TR to being admitted to hospital (mediators) i.e. variables that are affected by the intervention itself. The following variables are expected to be included in the propensity score analysis: age, sex, ethnicity, marital status, sexual orientation, deprivation score, relevant diagnosis (intellectual disability with/without autism, or autism without intellectual disability), pre-existing psychiatric diagnoses.

#### Stage 2: Difference-in-Differences (DiD) analysis

The DiD method compares changes in the prevalence of people admitted to hospital over time, contrasting the group who have a C(E)TR and the group who do not have a C(E)TR and attributes the “difference-in-differences” to the effect of the intervention.^14,15^ Here, we will apply the DiD model to examine the causal effect of a C(E)TR on the prevalence of psychiatric hospital inpatient status at any point over the 6-month period following the C(E)TR. Propensity scores will be included as weightings in the DiD analysis, following the method proposed in

Stuart (2014). The estimand will be the Average effect of the Treatment on the Treated (ATT). This is the average effect of receiving a C(E)TR on inpatient status in the 6 months after the C(E)TR. The data to be entered into the DiD model is considered panel data - repeated-measures data from the same patients before and after the index date - thereby strengthening the causal claim. If the DiD model described above fails to converge or if important assumptions are not met (Supplementary Text), we will use the propensity scores in a logistic regression analysis, with 6 month post-index date hospital inpatient status (yes/no) as the dependent variable and group (C(E)TR intervention/control) as an independent variable.

#### Sensitivity analyses

Sensitivity analyses will be carried out on the causal framework to probe the robustness of the applied model. During the C(E)TR identification process, we will identify patients for whom a community C(E)TR was suggested where there was no evidence that a C(E)TR did in fact occur. These are to be included in the pool of control patients in our main analysis. Exploratory sensitivity analyses will: 1) re-assign these patients to the C(E)TR group, and use the date in the record when a C(E)TR was suggested (or soon thereafter) as the index date, and/or 2) directly compare the main C(E)TR group with the ‘C(E)TR suggested’ group.

Some patients may have multiple community C(E)TRs while under follow-up. These will be treated as separate events in the main analysis (if the C(E)TRs are at least 6 months apart). Patients who have multiple C(E)TRs may have a different profile of baseline characteristics than those who receive only one. This may lead to pre-intervention differences in the C(E)TR versus no C(E)TR groups that cannot be sufficiently controlled for using the propensity scoring method. A sensitivity analysis will be performed on the sub-group of patients who only had one C(E)TR (versus no C(E)TR), improving sensitivity to the causal effect of a single C(E)TR.

### Sub-study 2 – Community cohort

**PICO framework** (Table 1)

#### Index dates

The index date will be defined as the date of admission to psychiatric hospital.

#### Outcomes

Outcome measures are the length of hospital stay (number of days), and clinical and functional improvement whilst in hospital. The Health of the Nation Outcome Scales (HoNOS) is a clinician-rated clinical and social functioning assessment scale with 12 items, each rated between 0 (no problem) and 4 (very severe problem).^16^ It has good psychometric properties.^17,18^ The HoNOS is routinely recorded by NHS staff at intervals and at points of care transition (e.g. hospital admission and discharge).

### Inclusion and exclusion criteria

All eligible patients who were admitted to psychiatric hospital for at least 14 days may be included in this study. We will exclude people who were not open to a SLaM community team for at least 6 months prior to their admission as these people may have a high degree of missing data and may not have had an opportunity for a community C(E)TR before their admission or might represent new transfers from other NHS Trusts. We will also exclude ‘re-admissions’, defined as an admission occurring <6 months after the discharge date of a previous admission.

### Identifying cases and controls

Using the review of clinical records of eligible patients previously described, we will measure whether each patient had a community C(E)TR within six months prior to their psychiatric hospital admission. We will form two groups; those who had a pre-admission community C(E)TR, and those who did not.

### Statistical analysis

#### Baseline descriptives

We will report the socio-demographic variables at admission (index date) and compare variables between those who had a pre-admission community C(E)TR and those who did not (Supplementary Tables 1 and 2). As described in sub-study 1, frequencies and percentages will be obtained for categorical variables and mean (SD) or median (IQR) for continuous and count variables, according to their distributions. Data will be also be presented separately for 1) people with intellectual disability and/or autism, and 2) autistic people without intellectual disability. Demographic and clinical characteristics will be compared using standard statistical comparisons for categorical (Chi square) and continuous variables (t-test or Kruskall-Wallis).

#### Pre-admission clinical and service use analysis

We will report clinical and service use variables for the 6 months before hospital admission and compare variables between those who had a pre-admission community C(E)TR and those who did not. This will also be implemented using standard statistical comparisons for categorical (Chi square) and continuous variables (t-test or Kruskall-Wallis).

#### Time-to-event analysis

Time-to-discharge (length of hospital admission) will be analysed using the Kaplan-Meier estimator.^19^ A Cox’s proportional hazard model will be used to estimate the hazard ratio for discharge,^20^ comparing those who received a C(E)TR to those who did not receive a C(E)TR for a follow-up period of six months after admission. This analysis will be adjusted for C(E)TR status prior to admission, age at admission, sex, ethnicity, and relevant diagnosis at admission (intellectual disability with/without autism, or autism alone). Patients who die within 6-months of admission will be censored at the date of death, and patients who remain in hospital will be censored at the end of the 6-month period.

#### Improvement in clinical and social functioning during hospital admission

Although we expect variation in how frequently HoNOS scores are collected and some missing data, we will attempt to extract HoNOS scores at two time-points of a patient’s psychiatric admission; the closest HoNOS score recorded around the admission date (+/- a maximum of 3 months) and the closest recorded HoNOS score to the discharge date (+/- a maximum of 3 months). A linear regression model will be carried out with the difference in total HoNOS score as the dependent variable. C(E)TR group will be the main independent variable of interest, with baseline HoNOS included as a covariate. Age, sex, and ethnicity will be included as confounders. We want to understand if having a community C(E)TR before hospital admission is associated with greater improvements whilst in hospital. As variations of HoNOS are applied across different populations, we will carry out this analysis on the version that has the least missing data.

### Missing data

The design of the study ensures that the intervention and outcome are fully-observed, however owing to the nature of the data source, we anticipate non-trivial proportions of missing confounder data. We will report the proportion and frequency of missing values for covariates. We will consider the missingness pattern approach, a technique which, if key assumptions are not violated, retains all people in the analysis. We will undertake various sensitivity analyses to explore whether our conclusions are affected when the assumption for the missing pattern approach is violated.^21^ A similar approach will be applied to the economic analysis, where missing data will be addressed using multiple imputation by chained equations with predictive mean matching.^22^

### Extensions of sub-study 1 and sub-study 2

#### Economic analysis

A partial economic evaluation will assess the cost impact of community C(E)TRs on secondary healthcare use. The primary outcome will be total secondary healthcare cost per patient, comparing those who received a community C(E)TR with those who did not. Service use data will be derived from CRIS and HES. Database accessibility and relevant resource use variables will be reviewed to determine feasibility. If both CRIS and HES are used, discrepancies for the same patient will be checked manually. Due to the linkage, the end date for the HES data extraction will be 31 March 2024. For HES data, the HRG Grouper and Spell Converter will be used to classify activity and define spells of care. Unit costs will be based on NHS Reference Costs and PSSRU.^23^ Mean cost differences and 95% confidence intervals will be estimated using non-parametric bootstrap regression (5,000 replications), adjusting for baseline costs and covariates from the main analysis. Service-specific utilisation will be summarised descriptively without formal comparisons, to focus on total cost and avoid multiple testing issues.

#### Analysis of a 12-month horizon

We will re-run the analyses for both sub-studies across a12-month horizon, to extend the results of the reported 6-month horizon and to capture potential impacts of community C(E)TRs across a longer time-frame.

#### Replication

We will replicate both sub-studies in the EHR of North London Foundation Trust (NLFT). NLFT provides a similar range of secondary mental healthcare services to SLaM (the primary site of this study). This will test the generalizability of the findings, by examining health records in another region and for a different demographic population. Under ethical and data governance approvals, it is not possible to pool data originating from SLaM and NLFT.

### Software

Statistical tests, data pre-processing, visualisations and diagnostics will be undertaken using R. Propensity score weighting and difference-in-differences models will be implemented in Stata.

### Patient and public involvement and engagement

The study received local approval from the SLaM CRIS Oversight Committee, chaired by a patient representative, which reviews and approves applications to extract and analyse data using CRIS. Progress reports are submitted to the Oversight Committee periodically.

PPI representatives and experts-by-experience are co-investigators on the grant that funds this research and contributed to the research questions and overall design of the study. They convene and run regular PPI group meetings with children and young people with intellectual disability and/or who are autistic, autistic adults, adults with intellectual disability, and family carers. PPI group members will assist with interpretation of the findings, suggesting clinical implications, and dissemination activities including any recommendations that arise as a result of the work.

### Ethics and dissemination

Access to the data and study approval was granted by the CRIS Oversight Committee (chaired by a patient representative) in accordance with the SLaM CRIS overarching ethical approvals for research use of extracted clinical data (Oxfordshire Research Ethics Committee C, ref:18/SC/0372). The NLFT CRIS database has been granted ethical approval from the East of England Research Ethics Service Committee - Cambridge Central, ref: 19/EE/0210), We will publish our findings in an open access peer-reviewed academic publication and present the results at relevant conference presentations. A full report detailing the work will be provided to the Funder. We will work with our PPI groups to ensure that the findings are available in accessible formats, including as easy read documents, so that they can be shared with people with intellectual disability and autistic people.

### Data management and confidentiality

To comply with all necessary data protection regulations, EHR data will be stored and analysed using encrypted workstations and virtual machines hosted in a UK-based Microsoft Azure datacentre. Data linkage between CRIS and HES datasets will be conducted in a secure environment. Research team members will not have access to any identifiable patient data. De-identified aggregate results from the dataset will be included in a peer-reviewed academic publication.

## Author contributions

RS and AA conceived the project and acquired funding. BM, LB, BC, and RS planned the analyses with input from RSt, JD, and AA. BM and RS wrote the original draft with subsequent review and editing by all authors. All authors approved the final version and jointly take responsibility for the decision to submit the manuscript for publication.

## Funding statement

This study is funded by the NIHR Health Services & Delivery Research (ref: NIHR158490).

## Competing interests statement

None declared.

## Data Availability

No data are available.

## SUPPLEMENTARY TEXT

### Causal model and assumptions

We will explicitly invoke knowledge-based causal assumptions needed to bridge the statistical interpretation of the intervention effect to a causal interpretation. These are the assumptions of consistency, positivity, pre-intervention parallel trends, stable unit treatment values, and ignorability, described as follows:

i. Consistency: we assume that the intervention, in this case, the community C(E)TR, is clearly defined, that there are no different versions of the treatment, and that the observed outcomes are consistent with the potential outcomes.
ii. Positivity: we assume that there is some chance (i.e. non-zero probability) of individuals in the study receiving the treatment. That is, there are no deterministic rules which force some individuals to either receive or not receive a C(E)TR.
iii. Pre-intervention parallel trends assumption: we assume that the time trends in the outcome are the same for both the patients who received a C(E)TR and did not receive a C(E)TR. We also assume that exogenous shocks (such as the start of the COVID-19 pandemic) impact both groups equally in the post-intervention period.
iv. Stable unit treatment values assumption: also known as no interference, we assume that the treatment of one person does not affect the outcome of other individuals.
v. Ignorability assumption: we assume that there are no unobserved differences between the treatment and control groups, conditional on the observed covariates.

When interpretating our findings, we will consider the extent to which they violate these assumptions. We will also examine the use of time-varying co-variates in our models; since C(E)TRs are convened when a service user is considered at high risk of being admitted to psychiatric hospital, there is high overlap between mental health and service use variables that lead to a C(E)TR and also lead to the need for hospitalization. Furthermore, any actions or recommendations made at the C(E)TR are specifically tailored to reduce the risk of hospitalization. This leads to a high overlap between variables that are associated with both receiving a C(E)TR and hospitalisation (i.e, they are confounding variables), and may also lie on the causal pathway (i.e. they are mediators) between having a C(E)TR and being admitted to hospital. This presents unique considerations, as only time-varying co-variates that contribute to both hospital admission and the C(E)TR itself should be included in our models (i.e. confounders only). We will therefore not include any time-varying covariates that we posit to be both mediators and confounders in our statistical models.

---

**SUPPLEMENTARY TABLE 1.**
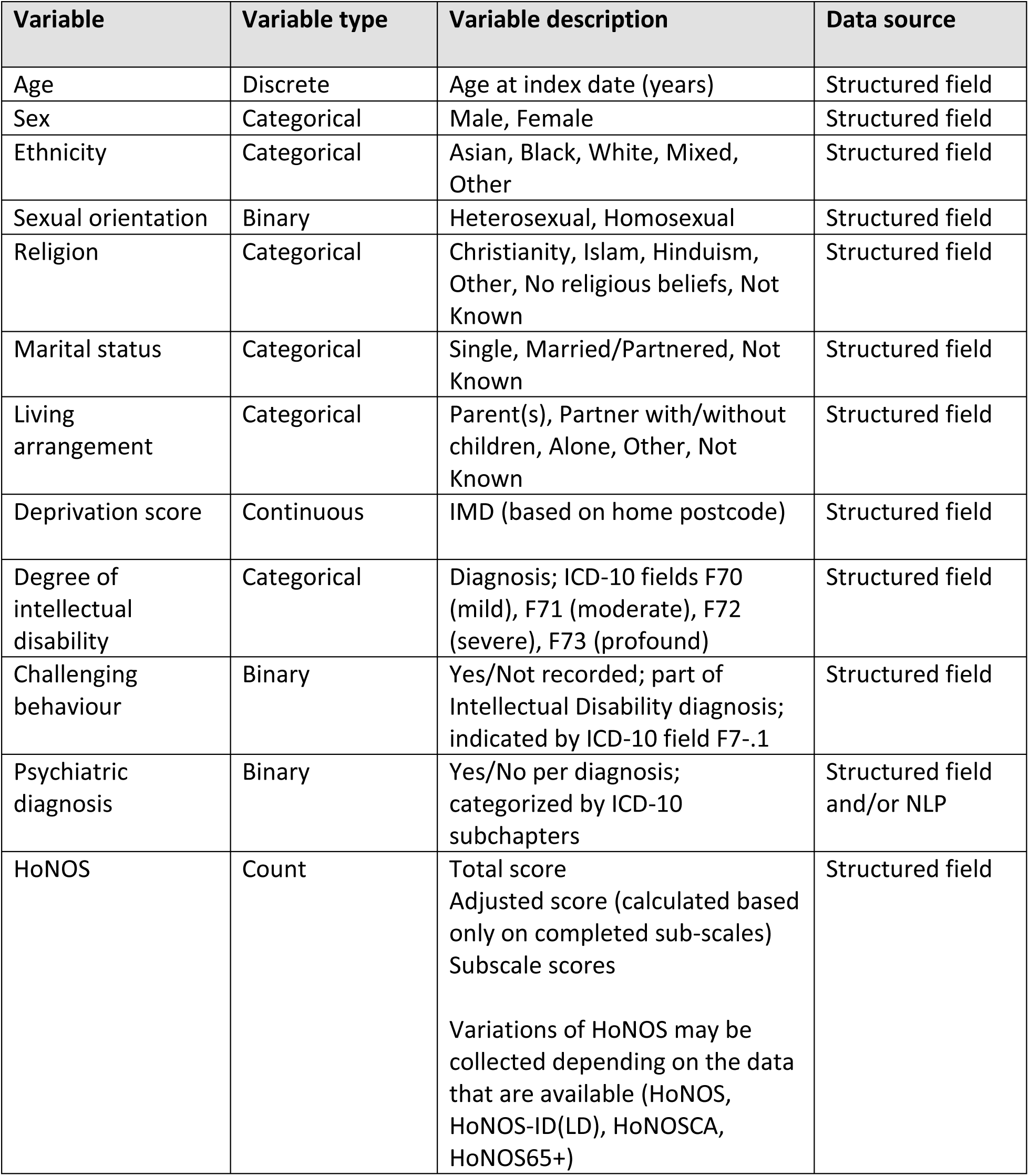
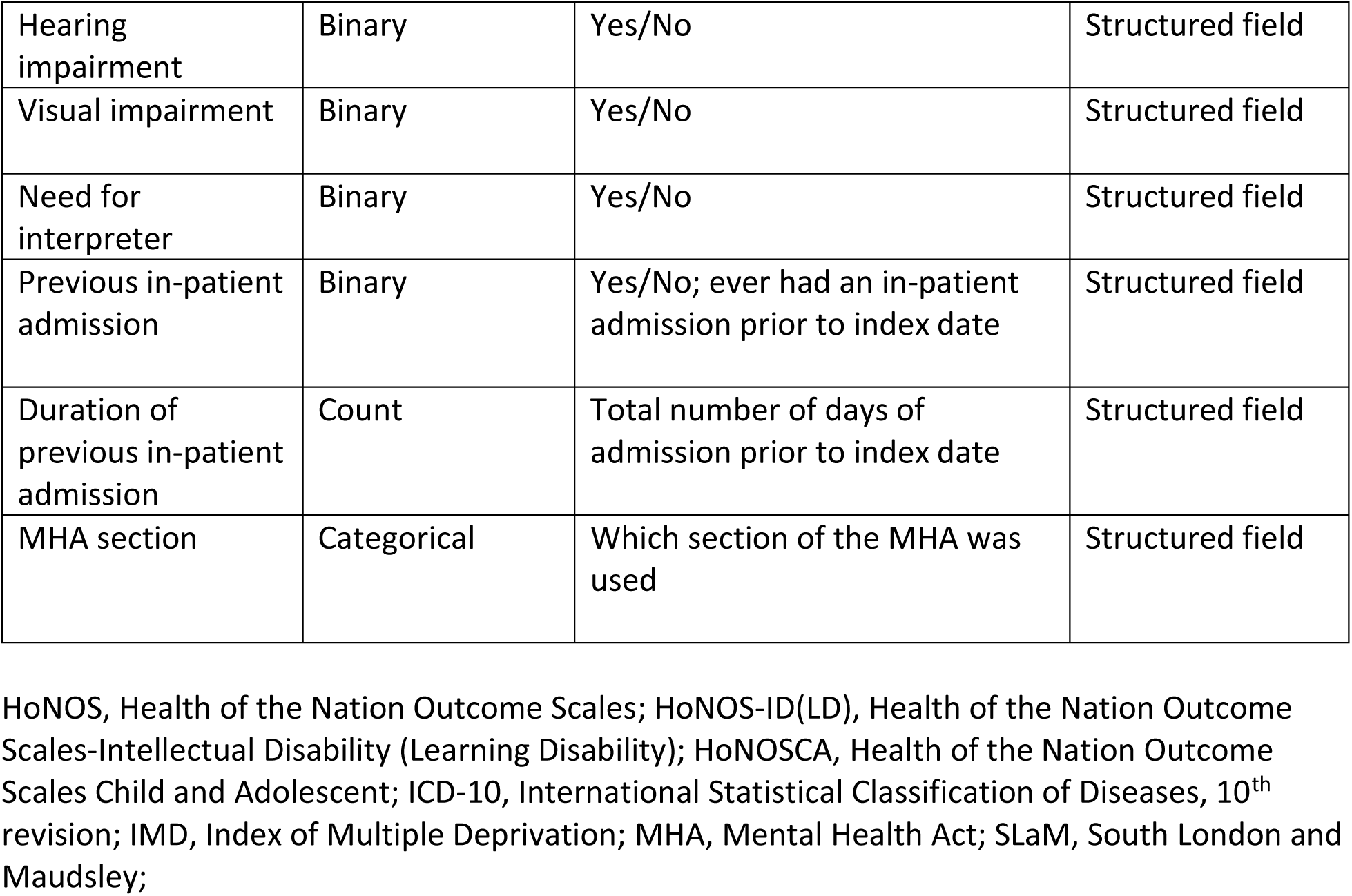
Baseline and time-varying socio-demographic and clinical variables.

Baseline and time-varying socio-demographic and clinical variables
| Variable | Variable type | Variable description | Data source |
| --- | --- | --- | --- |
| Age | Discrete | Age at index date (years) | Structured field |
| Sex | Categorical | Male, Female | Structured field |
| Ethnicity | Categorical | Asian, Black, White, Mixed, Other | Structured field |
| Sexual orientation | Binary | Heterosexual, Homosexual | Structured field |
| Religion | Categorical | Christianity, Islam, Hinduism, Other, No religious beliefs, Not Known | Structured field |
| Marital status | Categorical | Single, Married/Partnered, Not Known | Structured field |
| Living arrangement | Categorical | Parent(s), Partner with/without children, Alone, Other, Not Known | Structured field |
| Deprivation score | Continuous | IMD (based on home postcode) | Structured field |
| Degree of intellectual disability | Categorical | Diagnosis; ICD-10 fields F70 (mild), F71 (moderate), F72 (severe), F73 (profound) | Structured field |
| Challenging behaviour | Binary | Yes/Not recorded; part of Intellectual Disability diagnosis; indicated by ICD-10 field F7-.1 | Structured field |
| Psychiatric diagnosis | Binary | Yes/No per diagnosis; categorized by ICD-10 subchapters | Structured field and/or NLP |
| HoNOS | Count | Total score<br>Adjusted score (calculated based only on completed sub-scales)<br>Subscale scores<br>Variations of HoNOS may be collected depending on the data that are available (HoNOS, HoNOS-ID(LD), HoNOSCA, HoNOS65+) | Structured field |
| Hearing impairment | Binary | Yes/No | Structured field |
| Visual impairment | Binary | Yes/No | Structured field |
| Need for interpreter | Binary | Yes/No | Structured field |
| Previous in-patient admission | Binary | Yes/No; ever had an in-patient admission prior to index date | Structured field |
| Duration of previous in-patient admission | Count | Total number of days of admission prior to index date | Structured field |
| MHA section | Categorical | Which section of the MHA was used | Structured field |

HoNOS, Health of the Nation Outcome Scales; HoNOS-ID(LD), Health of the Nation Outcome Scales-Intellectual Disability (Learning Disability); HoNOSCA, Health of the Nation Outcome Scales Child and Adolescent; ICD-10, International Statistical Classification of Diseases, 10<sup>th</sup> revision; IMD, Index of Multiple Deprivation; MHA, Mental Health Act; SLaM, South London and Maudsley;

**SUPPLEMENTARY TABLE 2.**
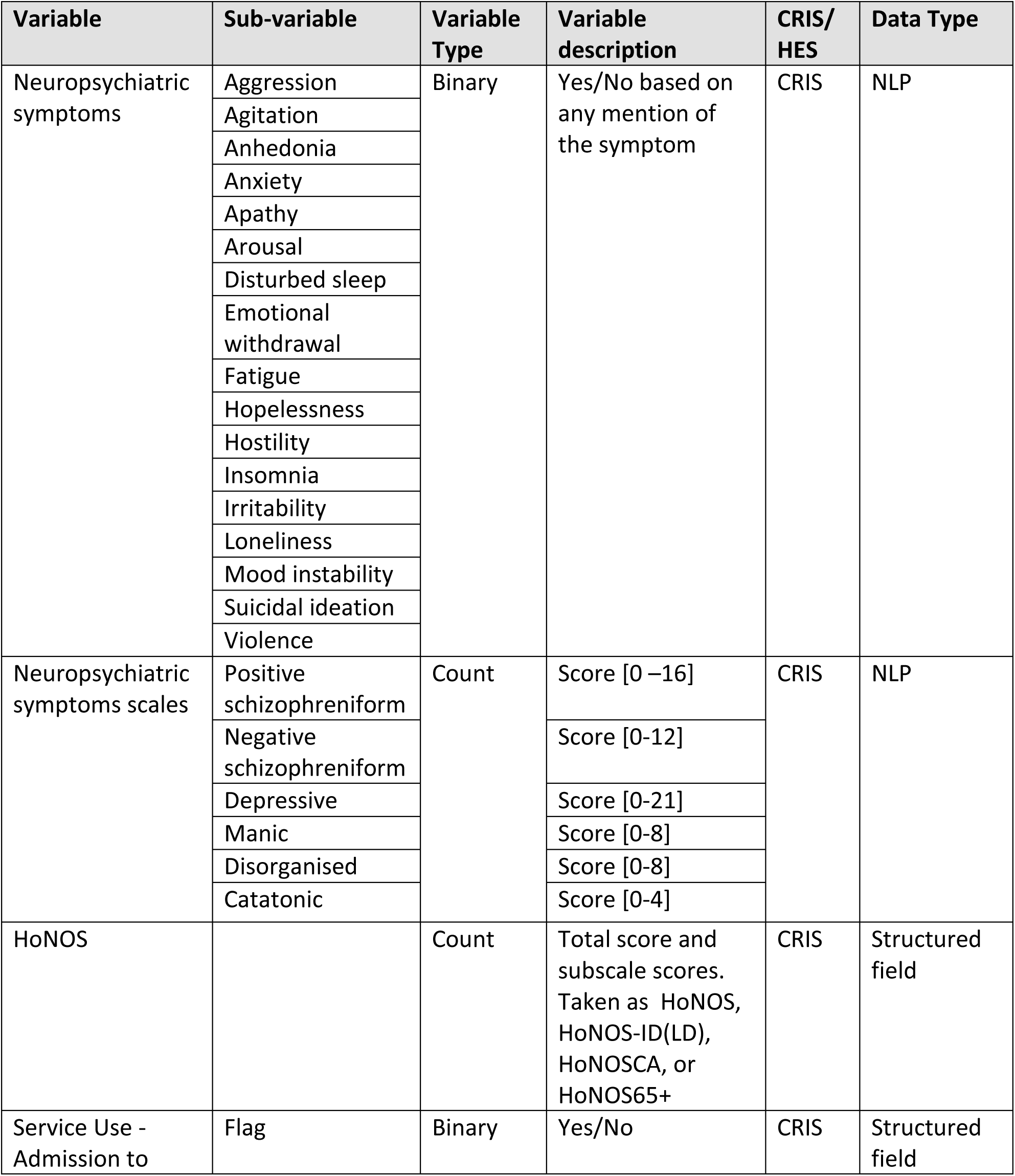
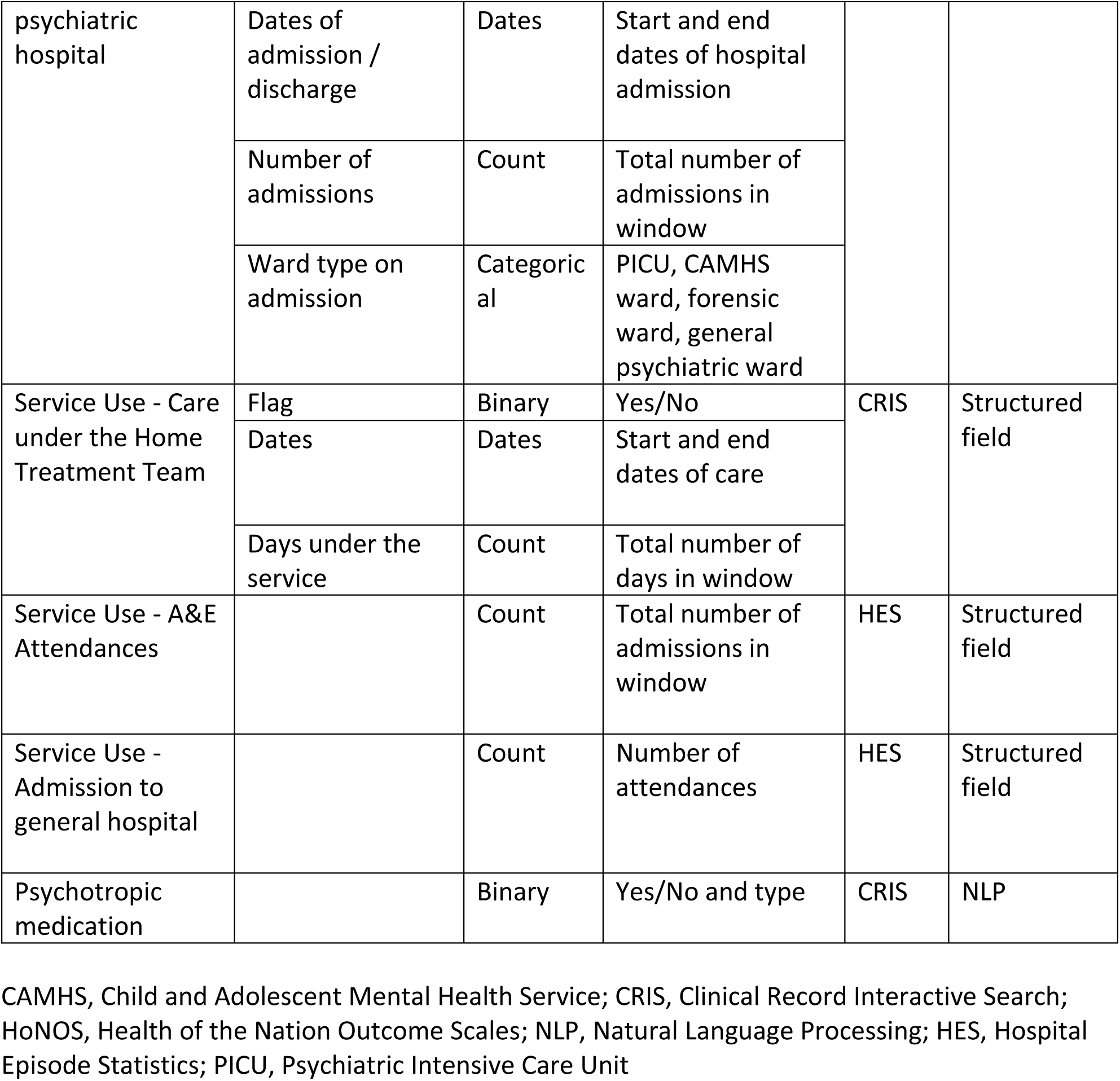
Pre-/post-index date variables for sub-study 1 and sub-study 2.

## Notes

### Competing Interest Statement

The authors have declared no competing interest.

### Author Declarations

Access to the data and study approval was granted by the CRIS Oversight Committee (chaired by a patient representative) in accordance with the SLaM CRIS overarching ethical approvals for research use of extracted clinical data (Oxfordshire Research Ethics Committee C, ref:18/SC/0372). The NLFT CRIS database has been granted ethical approval from the East of England Research Ethics Service Committee - Cambridge Central, ref: 19/EE/0210),

